## Supplementary Information for "Drivers and impact of the early silent invasion of SARS-CoV-2 Alpha"

### This PDF file includes :

- Supporting text
- Supplementary Fig. 1 to 5
- Supplementary Tables 1 to 5
- SI References

### Supporting information Text

#### *Arrival times, number of passengers and sequencing coverage*

Assuming exponential increase of cases in the origin country, the time of first arrival with passenger flow $p$ is Gumbel distributed with mean proportional to $\log(p)$ [1]. Assuming collection coverage $s$ on top, the mean arrival time based on first collection scales as $\log(p) + \log(s)$ and "iso-arrival-time" lines are as $\log(p) + \log(s) = k$ (anti diagonals). We report such lines in Fig. 1B and note that this scaling is consistent with the data.

#### *International dissemination model: Sensitivity analysis*

In the sensitivity analysis we tested the following assumptions:

- Mean incubation period equal to 4 days
- Mean incubation period equal to 6 days
- Delays computed for each countries after the alert averaged over a sliding time window of 7 days
- Percentage of case detection outside the UK, $K_c$= 25%
- Flights from all England airports with a catchment population of 56 millions inhabitants
- No changepoint for the Alpha incidence exponential growth in the UK
- Two changepoints for the Alpha incidence exponential growth in the UK an 5 Nov 2020 and 2 Dec 2020

Supplementary Table 5 shows the results of the sensitivity analysis. For the baseline scenario and the sensitivity models tested we provide best estimates and some model predictions chosen as reference. Varying the parameters had little impact on the parameters estimated in the model. The number of countries with introduction before 31 Dec 2020 increased in the following cases: delays from collection to submission for Alpha computed for each country aggregated over 7 days, 25% detection of imported cases, no change of slope.

#### *Local dynamics in the USA at a finer spatial scale*

The analysis of Alpha local spread for the USA shows that this country is out of trend, with a high number of predicted Alpha cases compared with model estimates. Here, we carry out the comparison for two individual states, California and Florida, and for New York City - see sources of data reported in Supplementary Table 2. These locations were the port of entry of Alpha into the US, with early reported Alpha cases linked directly to the UK [2,3].

As for the USA as a whole, the autochthonous model A was fed with importation fluxes estimated from the international dissemination model and we compared the model-predicted number of Alpha cases with the empirical estimates. We present these results on Supplementary Fig. 4.

For California and New York City, the comparison between model and empirical estimates follows a trend similar to European countries. Florida registered a high proportion of Alpha cases [2]. Such a high level of Alpha circulation can be compatible with model predictions in a scenario of early Alpha introduction, i.e. introduction dates close to the lower bound of the range predicted by the model.

#### *Median seeding time*

In the reference case in which traveling fluxes are constant in time and $R_t$ is the same in the destination country as in the UK, the reference median date of seeding would fall halfway between the date of emergence and 31 Dec 2020 (see Material and Methods). The median seeding dates of active chains at the end of 2020 departed from this assumed scenario. In Fig. 4D, we found that there was a negative correlation between the overall reproduction ratio over the period and the difference between computed median seeding date and reference median seeding date, implying that lower transmissibility overall led to less success in early introductions. In Supplementary Fig. 5 we show that there was no significant correlation between the international traffic drop and the difference of the median seeding date with the reference date.

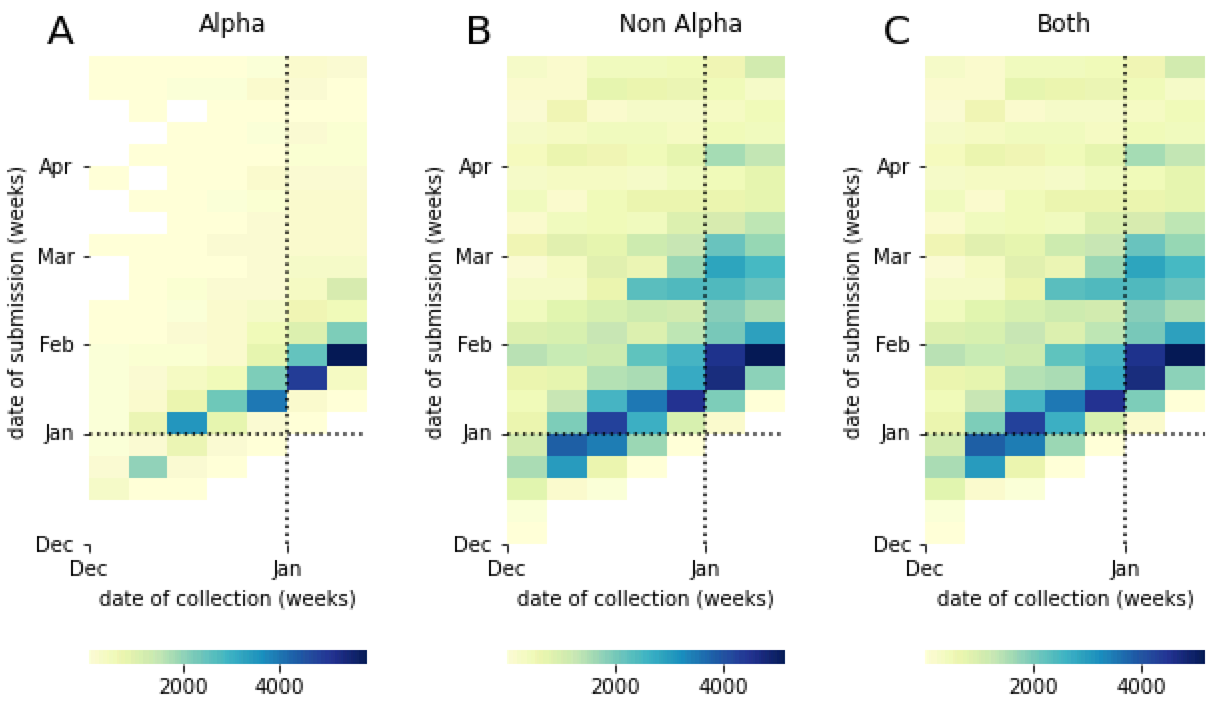


**Supplementary Fig 1. Occurrences of delays between collection and submission in time.** (**A**) Alpha variant.(**B**) Non Alpha variants. (**C**) Both.

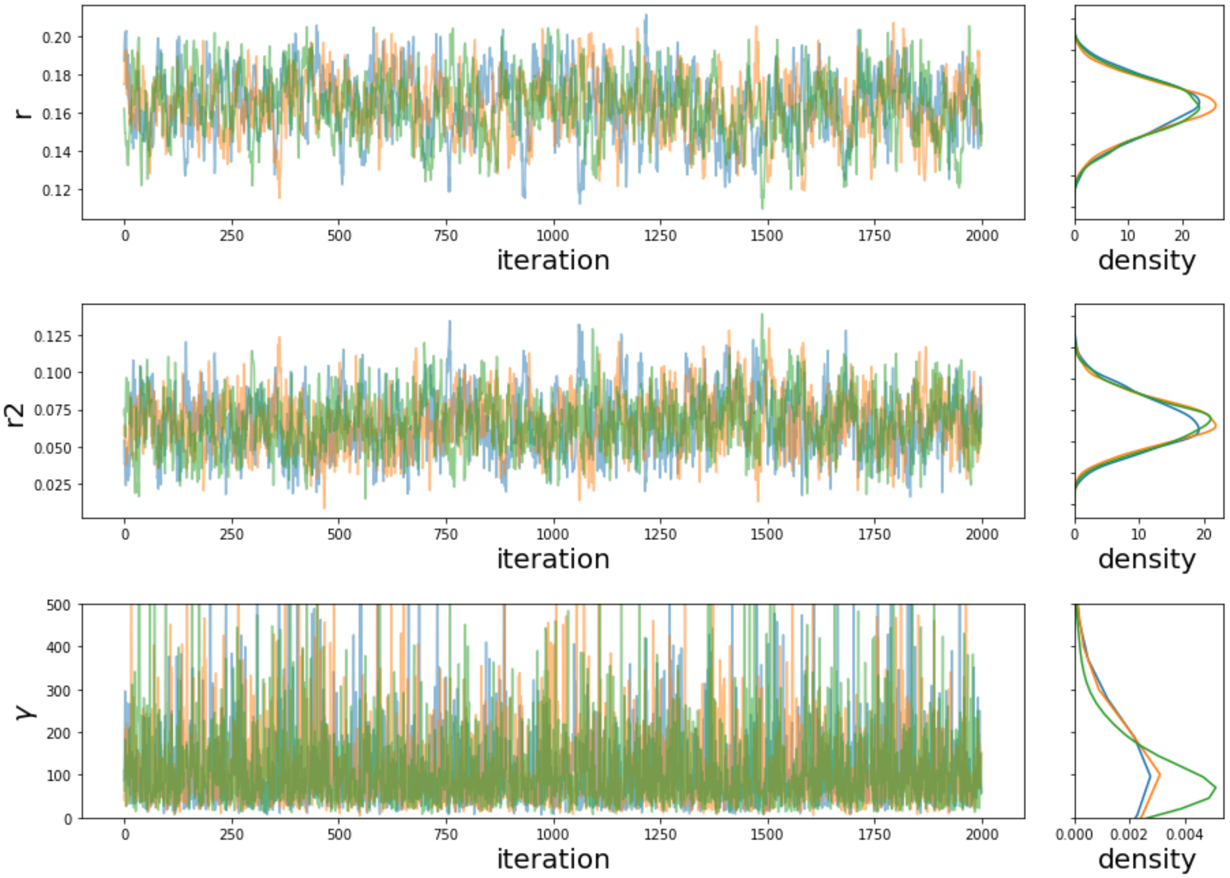


**Supplementary Fig. 2. MCMC convergence plot.** The fitted model has 2 exponential growth rates (r,r2) with changepoint on November 5th,2020. Three independent chains (red, green,blue) were run for 100000 iterations, with 50000 discarded as burn-in. Posterior samples were thinned 1 in 25.

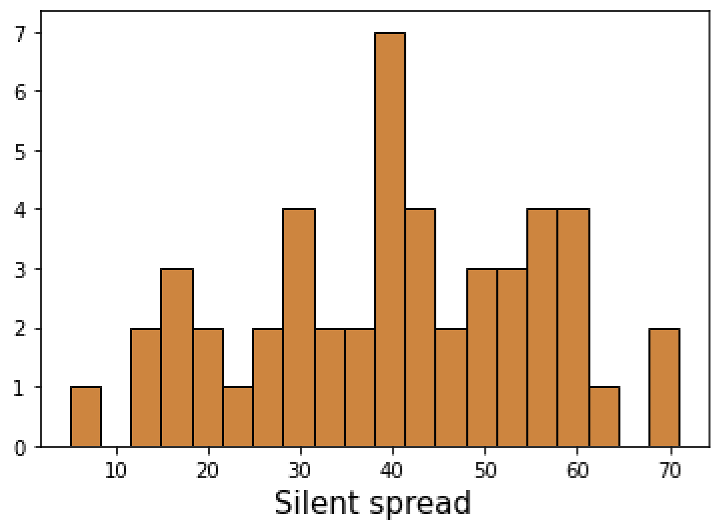


**Supplementary Fig. 3** Distribution of silent spread in days. Silent spread is computed as in Fig. 3D.

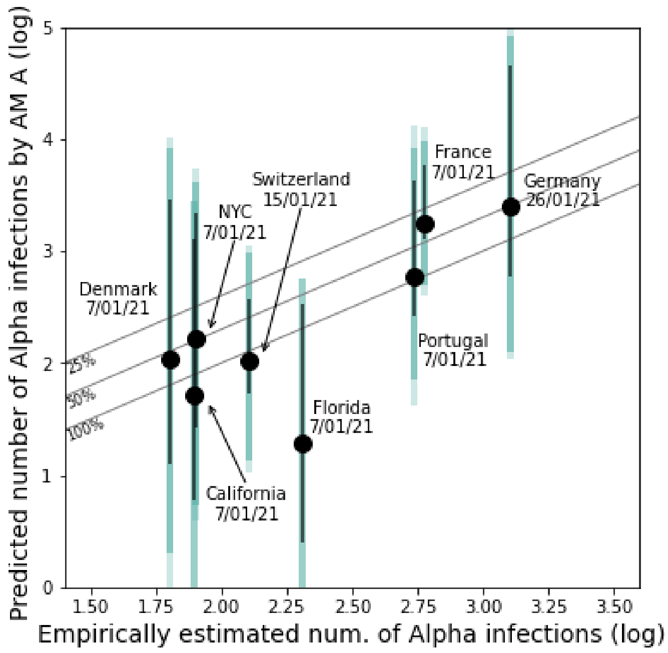


**Supplementary Fig. 4.** Model vs. empirical cases of Alpha as in Fig. 4 of the main paper. Here, the USA is replaced by California, New York City (NYC) and Florida to address spatial heterogeneity inside the USA.

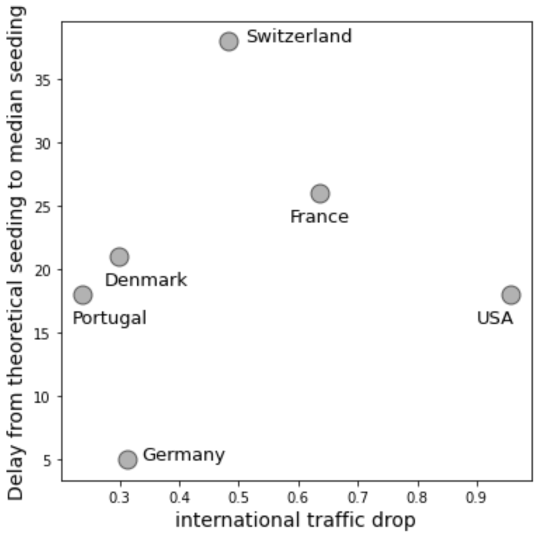


**Supplementary Fig. 5.** Difference between the median seeding date predicted by the autochthonous model A with the same quantity when $R_t$ is the same in all countries and traveling fluxes do not change in time, plotted against the international traffic drop. The international traffic drop is computed as the international traffic in Nov 2020 divided by the average of international traffic between Sep 2020 and Oct 2020. The Spearman correlation coefficient does not show a correlation between the two quantities (coefficient = 0.23, p-value = 0.66).

### Supplementary tables

| Country | Date of cases surveyed for Alpha | Daily number cases | Frequency Alpha | Computed number of Alpha infections | Source |
|---|---|---|---|---|---|
| France | 7 Jan 2021 | 18,004 | 0.033 | 594 | [4] |
| Portugal | 4-10 Jan 2021 | 8,062 (7/01/2021) | 0.068* | 548 | [5] |
| Germany | 23-29 Jan 2021 | 12,370 (26/01/2021) | 0.103 | 1,274 | [6] |
| Denmark | 4-10 Jan 2021 | 1,825 (7/01/2021) | 0.035 | 64 | [7] |
| Switzerland | 15 Jan 2021 | 2,204 | 0.058 | 128 | [8] |
| USA | 7 Jan 2021 | 248,566 | 0.0048 | 1,193 | [2] |

**Supplementary Table 1. Summary of data and assumptions used for the validation.** We assumed a fixed one-week delay between cases and infections. For example, infections of 31/12/2020 are detected on 07/01/2021. In Fig. 4A, we test the impact of a shorter (4 days) and higher (10 days) delay. France epidemiological report last accessed 26/05/2023. Germany report last accessed 26/05/2023. Denmark website last accessed 26/05/2023 (2 Mar 2021 version).

* At week 01 of 2021, 7.38% of cases were suspicions of Alpha, 92% of which are true Alpha.

| Country | Date of cases surveyed for Alpha | Daily number cases | Frequency Alpha | Computed number of Alpha infections | Source |
|---|---|---|---|---|---|
| California | 7 Jan 2021 | 43314 | 0.0018 | 78 | [2] |
| Florida | 7 Jan 2021 | 15939 | 0.0128 | 204 | [2] |
| NYC | 7 Jan 2021 | 5808 | 0.013738 | 80 | [9] |

**Supplementary Table 2.** Summary of data and assumptions used for the local spread analysis (as in Fig. 4A) for the additional states explored in Supplementary Fig. 4.

| Parameter | Description | Baseline value |
|---|---|---|
| $K_{UK}$ | Fraction of sampled Covid cases. | 0.25 |
| $K_c$ | Fraction of sampled imported Covid cases. | 0.5 |
| $e$ | Incubation period. | 5 days |
| $N$ | Population in the catchment area of London airports | 36M |
| $T_0$ | Beginning of the risk window for VOC emergence in the UK | August 1st, 2020 |
| $Tcp2$ | Date of change of the exponential transmission growth in the UK. | 5 Nov 2020 |

**Supplementary Table 3. Summary of the parameters values assumed in the international dissemination model.**

| Parameter | Description | Prior distribution |
|---|---|---|
| $r_1$ | Exponential growth rate in UK up to 5 Nov 2020 | Exp(0.1) |
| $r_2$ | Exponential growth rate in UK after 5 Nov 2020 | N(0,1) |
| $\gamma$ | Increasing factor of travelers sampling after 18 Dec 2020 | Exp(0.01) |

**Supplementary Table 4. Summary of the estimated parameters and their prior distribution.**

| Scenario | $r_1$ | $r_2$ | $\gamma$ | Predicted time of emergence in the UK | Median date of first introduction for France, and Denmark | #countries with introduction before 31 Dec 2020 |
|---|---|---|---|---|---|---|
| Baseline | 0.17 [0.14;0.20] | 0.055 [0.02;0.097] | 51.67 [12.43;310.11] | 08/09 [21/08;19/09] | 17/10 [18/09-03/11] - 05/11 [08/10-06/12] | 65 [52-73] |
| Incubation = 4 days | 0.17 [0.14;0.20] | 0.056 [0.024;0.096] | 51.51 [11.72;307.19] | 09/09 [21/08;18/09] | 17/10 [20/09-03/11] - 05/11 [09/10-07/12] | 65 [52-73] |
| Incubation = 6 days | 0.17 [0.14;0.20] | 0.056 [0.024;0.097] | 52.36 [11.89;301.70] | 06/09 [19/08;16/09] | 17/10 [19/09-03/11] - 05/11 [09/10-06/12] | 65 [52-73] |
| Delays computed by country, aggregated on 7 days After 18 Dec 2020 | 0.16 [0.11;0.19] | 0.086 [0.043;0.13] | 36.26 [7.60;186.32] | 02/09 [17/08;18/09] | 17/10 [20/09-09/11] - 09/11 [09/10-07/12] | 69 [60-73] |
| 25% detection of imported cases | 0.18 [0.15;0.21] | 0.058 [0.025;0.1] | 51.20 [11.53;300.63] | 06/09 [19/08;18/09] | 15/10 [19/09-31/10] - 31/10 [07/10-30/11] | 70 [61-73] |
| Air travel : flight from England | 0.17 [0.14;0.20] | 0.056 [0.025;0.097] | 73.13 [15.46;415.13] | 02/09 [17/08;18/09] | 19/10 [21/09-06/11] - 07/11 [10/10-08/12] | 64 [47-72] |
| No change of slope | 0.12 [0.11;0.13] | | 22.46 [6.75;94.21] | 30/08 [16/08;12/09] | 23/10 [14/09-13/11] - 24/11 [12/10-13/12] | 69 [59-73] |

| | | | | | | |
|---|---|---|---|---|---|---|
| 2 changes of slope : 11/5 and 12/2 | 0.16 [0.12;0.20] | r2 : 0.07 [0.026,0.15] r3 : 0.023 [0.022,0.48] | 61.32 [13.19;332.14] | 02/09 [17/08;17/09] | 18/10 [18/09-08/10] - 08/11 [09/10-07/12] | 65 [51-73] |

**Supplementary Table 5. Sensitivity analysis of the international dissemination model.**
